# Circulating tumor cell characterization of lung cancer brain metastasis in the cerebrospinal fluid through single-cell transcriptome analysis

**DOI:** 10.1101/2020.01.06.20016683

**Authors:** Haoyu Ruan, Yihang Zhou, Jie Shen, Yue Zhai, Ying Xu, Linyu Pi, RuoFan Huang, Kun Chen, Xiangyu Li, Weizhe Ma, Zhiyuan Wu, Xuan Deng, Xu Wang, Chao Zhang, Ming Guan

## Abstract

Metastatic lung cancer accounts for about half of the brain metastases (BM). Development of leptomeningeal metastases (LM) are becoming increasingly common, and its prognosis is still poor despite the advances in systemic and local approaches. Cytology analysis in the cerebrospinal fluid (CSF) remains the diagnostic gold standard. Although several previous studies performed in CSF have offered great promise for the diagnostics and therapeutics of LM, a comprehensive characterization of circulating tumor cells (CTCs) in CSF is still lacking. To fill this critical gap of lung adenocarcinoma LM (LUAD-LM), we analyzed the transcriptomes of 1,375 cells from 5 LUAD-LM patient and 3 control samples using single-cell RNA sequencing technology. We defined CSF-CTCs based on abundant expression of epithelial markers and genes with lung origin, as well as the enrichment of metabolic pathway and cell adhesion molecules, which are crucial for the survival and metastases of tumor cells. Elevated expression of *CEACAM6* and *SCGB3A2* was discovered in CSF-CTCs, which could serve as candidate biomarkers of LUAD-LM. We identified substantial heterogeneity in CSF-CTCs among LUAD-LM patients and within patient among individual cells. Cell-cycle gene expression profiles and the proportion of CTCs displaying mesenchymal and cancer stem cell properties also vary among patients. In addition, CSF-CTC transcriptome profiling identified one LM case as cancer of unknown primary site (CUP). Our results will shed light on the mechanism of LUAD-LM and provide a new direction of diagnostic test of LUAD-LM and CUP cases from CSF samples.

## Introduction

Lung cancer is the second most common cancer type in both men and women(1). Non-small cell lung cancer (NSCLC) is the main type of lung cancer, accounting for 85% of lung malignancies with a 5-year survival rate less than 15% (2). Histologically, NSCLC was further clarified into three subtypes: lung adenocarcinoma (LUAD), squamous-cell carcinoma and large cell carcinoma (3), among which LUAD is the most common histological subtype (4). Most lung cancer-associated morbidity results from metastases. Brain is the most common metastatic site of NSCLC and the incidence of brain metastases (BM) ranges from 22% to 54%, occurring at different stages of tumorigenesis, especially in advanced patients (5). Of all cancer patients with brain metastases, lung cancer is the primary tumor in 40 to 50% cases(6), which is the highest among all cancer types and equals all other primary cancer types combined (7). Leptomeningeal metastasis (LM) occurs in 3-5% of patients with advanced NSCLC and it is the most frequent in the LUAD subtype (8). Leptomeningeal metastases result from dissemination of cancer cells to both the leptomeninges (pia and arachnoid) and cerebrospinal fluid (CSF) compartment (9). In this study, we enrolled five LUAD patients with leptomeningeal metastases (LUAD-LM); four patients showed leptomeningeal enhancement by magnetic resonance imaging (MRI) of brain, and one patient only had tumor cells in CSF.

The diagnosis and monitoring of NSCLC-LM are based on neurological and radiographic imaging as well as CSF examinations. A positive CSF cytology result remains the gold standard for the diagnosis of NSCLC-LM and gadolinium-enhanced MRI of the brain and spine is the best imaging technique (10). In recent years, CSF liquid biopsy has attracted more attention. Tumor cells in CSF were considered as circulating tumor cells (CTCs) as CTCs in blood (11). The CellSearch technique (12), which utilizes immunomagnetic selection, identification and quantification of CTCs in the CSF, is more sensitive than the conventional cytology and MRI for the diagnosis of LM. Moreover, evaluation of CSF circulating free DNA (cfDNA) has the potential to facilitate and supplement LM diagnosis and the detection of genomic alterations can also inform targeted therapy (13). The aim of NSCLC-LM treatment is palliative including corticosteroids, chemotherapy, and radiotherapy which can extend survival to an average of less than one year (14). Intrathecal chemotherapy, molecularly targeted therapy and immunotherapy are also effective treatment to prolong survival (10). The development of new technologies and approaches have improved the diagnosis and therapy of NSCLC-LM, but the outcomes of brain metastases are still poor. To make breakthroughs in tackling the clinical challenge of brain metastasis, a comprehensive understanding of its mechanisms is needed. CSF sampling through lumbar puncture can help diagnose and monitor the progress of brain metastases, with the advantage of minimal risk compared to brain biopsy. By analyzing the transcriptome characteristics of tumor cells in CSF of NSCLC-LM patients, new biomarkers could be discovered for clinical diagnosis. In addition, knowledge of gene expression alterations in brain metastases compared to primary tumors will help develop novel targeted therapeutic approaches with improved efficacy.

CSF-CTCs was proposed as a novel tool for the diagnosis of LM (11). However, tumor cells are relatively rare in patient CSF samples, and ≥1 CSF-CTC/mL was defined as a cutoff for diagnosis (11). The traditional profiling technologies that measure tumor cells in bulk were confounded by the presence of normal lymphocytes and could not capture gene expression heterogeneity among tumor cells. Therefore, we investigated the transcriptome characteristics of CSF-CTCs by Smart-seq2 single-cell RNA sequencing (scRNA-seq). CSF-CTCs have not been investigated at the single-cell transcriptome level in leptomeningeal metastases. To date, there is only one CSF scRNA-seq study in the literature, which identified rare CNS immune cell subsets that may perpetuate neuronal injury during HIV infection (15). Here, we investigated the transcriptional profiles for more than one thousand CSF-CTCs from five LM patients, defined the CSF-CTC transcriptome profile and revealed intra-tumoral and inter-tumoral heterogeneity of patients for the first time.

## Results

### Cell composition of the cerebrospinal fluid at single-cell transcriptome level

To characterize the single-cell transcriptomes and the composition of CSF cells under healthy conditions, three samples (N1-N3) were used from patients who were screened for potential brain infections and diagnosed as normal (Supplementary Table 1). We sequenced the single-cell transcriptomes of 624 CSF cells using Smart-seq2 single-cell RNA-seq (scRNA-seq) technology (Figure 1A). Besides CSF cells, blood T cells and B cells were sorted and sequenced to establish the cell type transcriptome profiles to define the normal CSF cells composition. After quality filtering (see Methods), 207 normal CSF cells, 41 B cells and 41 T cells are clustered using t-distribution stochastic neighbor embedding (t-SNE) method (Figure 1B). On average, 803 expressed genes were detected per individual cell. We identified three clusters corresponding to B cells, T cells and monocytes defined by the expression patterns of lymphocyte markers (Figure 1C). The Blood-T and CSF-T cells are clustered together, indicating normal lymphocytes have similar expression profiles in different microenvironments. Normal CSF samples consist of 80.3% T cells and 19.7% monocytes. No B cells were found in CSF samples (Supplementary Table 2). Our result is consistent with previously known proportions in the textbook (16), which serves as a proof of principle of scRNA-seq in CSF samples.

**Figure 1.**
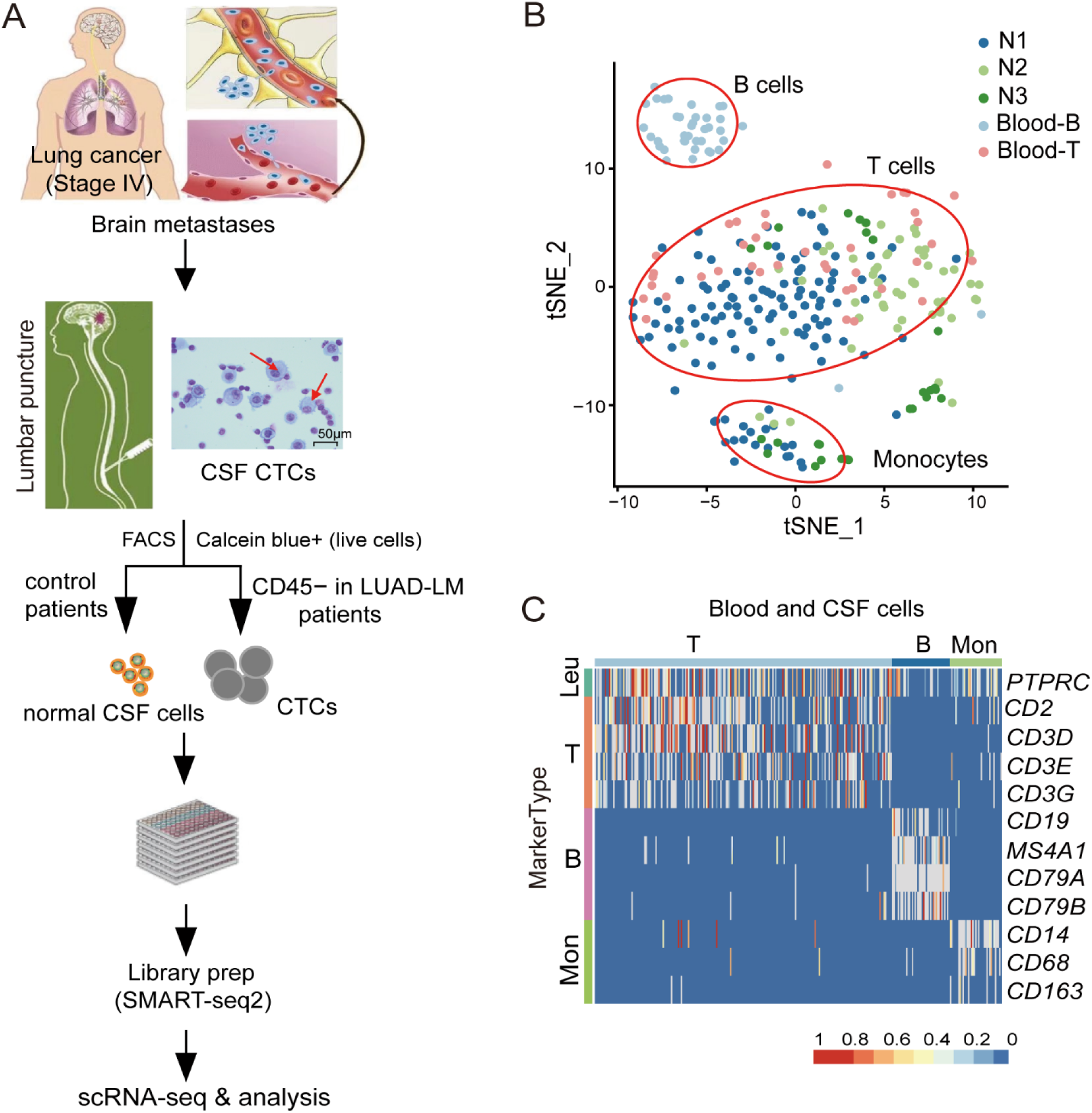
Isolation of cerebrospinal fluid (CSF) circulating tumor cells (CTCs) and characterization of normal CSF cell composition at single-cell level. (**A**) CSF-CTC isolation workflow. CSF samples were collected from lung adenocarcinoma (LUAD) leptomeningeal metastases (LM) patients. Under CSF cytology (magnification 400×; Wright’s stain), CTCs have a larger cell diameter (red arrows) than the smaller lymphocytes. Fluorescence-activated cell sorting (FACS) was used to sort individual live cells from LUAD-LM CSF samples, and CD45− cells were subsequently selected for scRNA-seq following the SMART-seq2 protocol. For control purpose, live cells from non-metastatic CSF samples (Normal, N1-N3) were processed using the same pipeline except for the cell selection. Illustration of LUAD-BM is modified from Fig.1 in Seyed H Ghaffari *et. al*. (**B**) t-distributed stochastic neighbor embedding (t-SNE) plot of cells from control CSF samples (N1, N2 and N3) and blood cells, showing the cell identity and gene expression correlation. (**C**) High-resolution heat map showing expression of selected marker genes in blood and CSF cells (Leu, Leukocytes; Mon, Monocytes).

### Identification and characterization of circulating tumor cells in the CSF of LUAD-LM patients

We developed an effective and highly reproducible protocol for cell isolation from the CSF samples of LUAD-LM patients (Figure 1A), and seven LM patients were enrolled in the scRNA-seq study (Supplementary Table 1). In total, 1,776 candidate CTCs from five LUAD-LM patients (P1, P2, P4, P6 and P7) were FACS sorted (CD45− and Calcein Blue AM+, Supplemental Figure 1) and sequenced, from which 1,152 cells with at least 600 covered genes in their transcriptomes are included in our analysis (Table 1), and these cells are clustered using the t-SNE method along with the three normal CSF samples N1-N3. The majority of patient CSF cells clustered according to the patient of origin, with the exception of 15 monocytes in P4 (Figure 2A). These patient CSF calls are candidate CTCs from the brain metastases. The clustering pattern is not driven by technical variability, because CSF samples collected from the same patient within a two-month time interval (P1-1 and P1-2) formed a single coherent cluster, despite that they have undergone independent cell sorting, library construction and sequencing (Figure 2A). We did not observe significant heterogeneity in mapping quality and gene coverage across patient samples (Supplemental Figure 2, A and B; Supplementary Table 1), suggesting the clustering is not due to technical artifact.

**Table 1.** Summary of cell type identity in scRNA-seq results of cancer patient CSF samples.

| Patient ID | Diagnostics | Cell selection |  | Number of sequenced cells | Number of QC filtered cells | T cells | Monocytes | CTCs |
| --- | --- | --- | --- | --- | --- | --- | --- | --- |
|  |  | Calcein Blue AM | CD45 |  |  |  |  |  |
| P1-1 | LUAD-LM | + | – | 168 | 99 | 0 | 0 | 99 |
| P1-2 | LUAD-LM | + | – | 360 | 280 | 0 | 0 | 280 |
| P2 | LUAD-LM | + | – | 192 | 130 | 0 | 0 | 130 |
| P4 | LUAD-LM | + | – | 288 | 157 | 0 | 15 | 142 |
| P6 | LUAD-LM | + | – | 480 | 281 | 0 | 0 | 281 |
| P7 | LUAD-LM | + | – | 288 | 205 | 0 | 0 | 205 |
| P3 | LUAD-LM | + | N/A | 96 | 50 | 0 | 50 | 0 |
| P8-1 | CUP-LM | + | – | 480 | 152 | 0 | 0 | 152 |
| P8-2 | CUP-LM | + | – | 816 | 343 | 28 | 19 | 296 |
| **Total** |  |  |  | **3,168** | **1,697** | **28** | **84** | **1,585** |

LUAD: lung adenocarcinoma; CUP: cancer of unknown primary site; LM: leptomeningeal metastases; Calcein Blue AM: labeling dye for live cells selection; CD45: protein tyrosine phosphatase receptor type C, marker for leukocytes; N/A: Not applicable.

**Figure 2.**
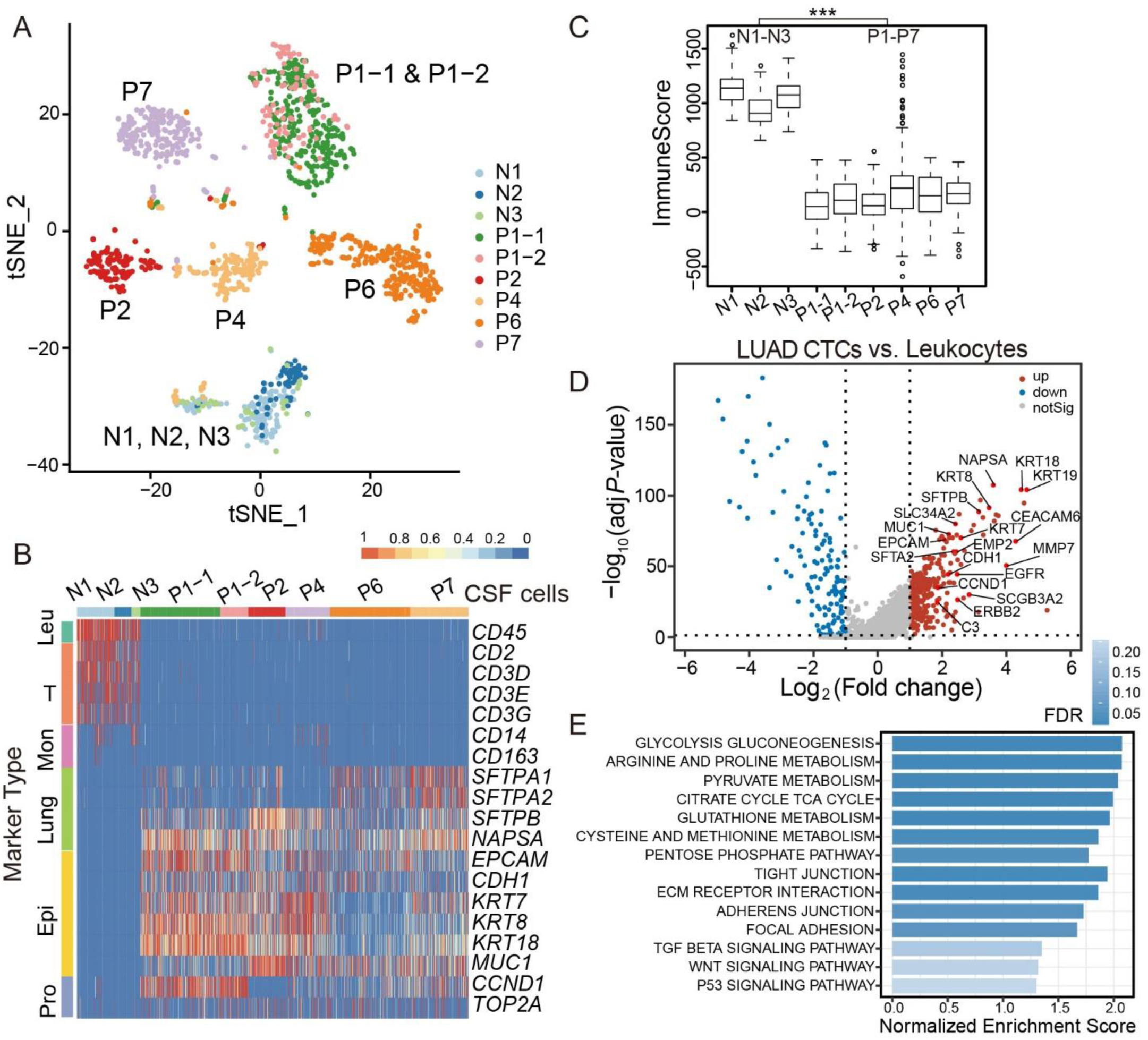
Characterization of LUAD-LM CSF-CTCs using single-cell transcriptome analysis. (**A**) t-SNE plot of gene expression clustering of three normal CSF samples (N1, N2 and N3) and five LUAD-LM CSF samples (P1, P3, P4, P6 and P7). Clusters are assigned according to cell identity and gene expression correlation. (**B**) Heatmap showing expression levels of selected marker genes in each sample (Leu, Leukocytes; Mon, Monocytes; Epi, Epithelial; Pro, Proliferation). (**C**) The immune signature of CSF cells quantified by the ImmuneScore computed from the ESTIMATE algorithm, showing the significant difference between the leukocyte group (***left***) and the CSF-CTC group (***Right***) (***: *P* < 0.001, Wilcoxon Rank-Sum test). (**D**) Volcano plot of differential gene expression profile between the CSF-CTCs in control and patients (adjusted *P*-value < 0.05; fold-change > 2). Gene names are labeled for selected upregulated genes in CSF-CTCs, including important internal reference and marker genes. (**E**) Significantly enriched KEGG pathways in LUAD CSF-CTCs compared to leukocytes by GSEA analysis (FDR: 0.05-0.20).

CSF-CTCs are larger than the CSF lymphocytes (Figure 1A). To determine whether CTCs could be separated by morphology without CD45− selection, live cells were isolated from another LUAD-LM patient (P3) merely based on cell morphology. Single-cell transcriptome profiling revealed 100% of collected cells are monocytes (Supplemental Figure 2C), indicating the CD45 negative selection step is necessary for the successful isolation of CSF-CTCs.

At molecular level, we define CSF-CTCs as non-immune cells with transcriptome signatures for lung adenocarcinoma markers, epithelial markers and proliferation markers (Figure 2B and Supplemental Figure 3). Based on the criteria, 100% cells in P1, P2, P6 and P7 CSF samples were CTCs and 90.4% (142/157) of P4 CSF cells are CTCs (Table 1). Compared to control CSF samples, patient CSF samples lack expression from immune cell markers (Figure 2B) and have a significantly lower mean ImmuneScore(17) (127 vs 1,703, *P* < 0.001, Wilcoxon Rank-Sum test; Figure 2C). Epithelial cell markers, including *EPCAM, CDH1, KRT7, KRT8, KRT18* and *MUC1*, are expressed in CTCs, suggesting their epithelial origin (Figure 2B and Supplemental Figure 3). Lung adenocarcinoma markers *SFTPA1, SFTPA2, SFTA2, SFTPB* and *NAPSA* are also expressed in most CTCs, indicating that they are originated from primary lung cancer tumor cells (Figure 2b and Supplemental Figure 3). CTCs also have expression of common proliferation markers *CCND1* and *TOP2A* (Figure 2B and Supplemental Figure 3).

### Transcriptome signatures of CSF-CTCs in LUAD-LM patients

290 gene are significantly upregulated in patients CSF-CTCs compared to normal CSF cells (adjusted *P*-value *P*-adj. < 0.05 and log2 fold-change logFC > 1; Figure 2d). Among these genes, *CEACAM6* (*P*-adj. = 1.23 × 10^−76^; logFC = 6.22) is carcinoembryonic antigen cell-adhesion molecule and a biomarker for mucinous adenocarcinoma including colon, pancreas, breast, ovary and lung malignancies. Overexpression of *CEACAM6* has been shown to associate with poor prognosis due to its roles in cellular invasiveness, resistance to anoikis and metastatic potential(18). *SCGB3A2*, another marker for pulmonary carcinoma (19), is also significantly upregulated (*P*-adj. = 4.46 × 10^−33^; logFC = 3.06). *SCGB3A2* is a member of the secretoglobin (SCGB) gene superfamily mainly found in bronchial epithelial cells. It is a growth factor during fetal lung development (20) with anti-inflammatory function in the lung (21). As secreted proteins, elevated expression of *CEACAM6* and *SCGB3A2* have great potential in developing CSF immunoassay for LUAD-BM diagnosis.

Energy metabolism and cell adhesion pathways are significantly enriched in CSF-CTC transcriptomes (FDR < 0.05, Figure 2E). Glycolysis gluconeogenesis and pentose phosphate pathways are critical for tumor growth and the energy demand in the brain. The up-regulated cell adhesion pathways consist of tight junction, extracellular matrix receptor interaction and adhesion functions, indicating that CSF-CTCs possessed a higher adhesion strength, crucial for essential functions such as survival, proliferation, migration, and the ability to maneuver through the capillary-sized vessels to a new location (22, 23). *MMP7* (*P*-adj. = 2.94 × 10^−49^; logFC = 3.99), matrix metalloproteinase 7, is one such molecule expressed in epithelial cells, promoting carcinogenesis through induction of epithelial-to-mesenchymal transition (EMT) (24). Secreted MMPs can disrupt the basement membrane and facilitate the metastases to other sites (25). MMPs can also open the blood-brain barrier by degrading tight junctions proteins (26).

Recently the complement system proteins have shown to be involved in malignant epithelial cells. C3 (*P*-adj. = 8.13 × 10^−33^; logFC = 2.19) activates the C3a receptor in the choroid plexus epithelium to disrupt the blood-CSF barrier, allowing plasma amphiregulin and other mitogens to enter the CSF and promote cancer cell growth (27). In summary, CSF-CTCs have unique gene expression profiles with full capacity of cancer and metastases related functions. The single-cell transcriptome characteristics are reassuring that the patient CSF cells we isolated are indeed CSF-CTCs.

### Spacial and gene expression heterogeneities of LUAD-LM tumors

LM often occur at different brain locations, resulting in spacial heterogeneity of the metastatic tumors. We examined the five LUAD-LM patients and this is exactly the case (Figure 3A). To investigate gene expression heterogeneities at the single-cell level, we quantified pairwise correlations between the expression profiles of 967 single-CTC transcriptomes from the five LUAD-LM samples (Figure 3B), and discovered significant heterogeneity both among different patients and within individual patient among CSF cells (correlation coefficients ranging from −0.057 to 0.829). Inter-tumor heterogeneity is significantly greater than the intra-tumoral heterogeneity (mean correlation coefficient −0.009 vs. 0.029, *P*-value < 2.2e-16, Wilcoxon Rank-Sum test, Figure 3, B and C). To compare with primary tumors, we utilized single-cell expression data from human non-small cell lung cancer cell line H358, and human lung adenocarcinoma patient-derived xenograft (PDX) MBT15 and PT45 (28). The cell-to-cell correlations within individual primary tumor samples are significantly higher than that within individual patient CSF samples (mean correlation coefficient 0.092 vs. 0.029, *P*-value < 2.2e-16, Wilcoxon Rank-Sum test), indicating greater heterogeneity in clinic CSF samples (Figure 3, B and C).

**Figure 3.**
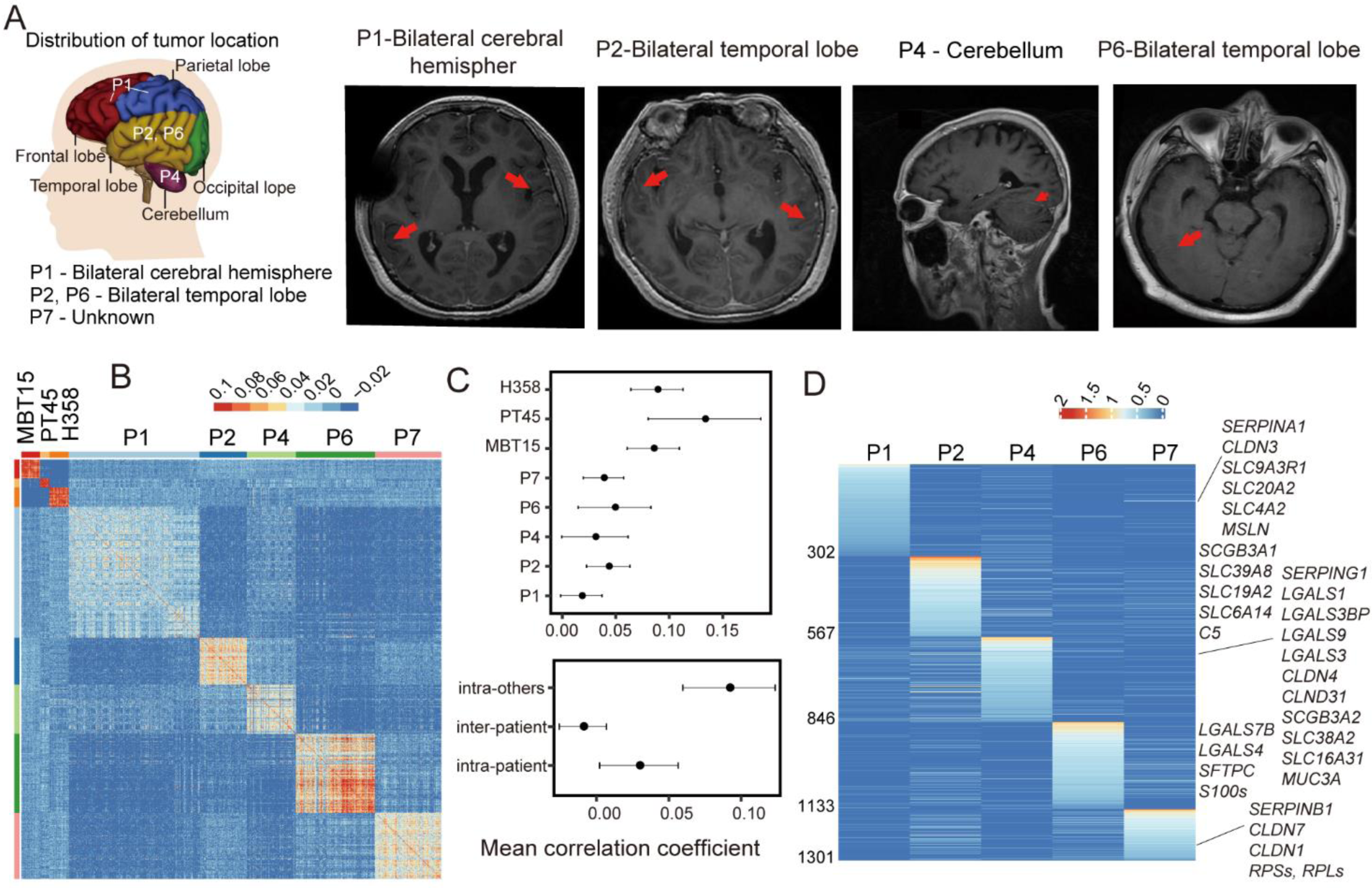
The heterogeneity of LUAD-LM CSF-CTCs among different patients and within individual patient. (**A**) Locations of leptomeningeal metastases in LUAD-LM patients reveal special heterogeneity. Brain MRI (Magnetic resonance imaging) diagnostic results were shown for patients P1, P2, P4 and P6, demonstrating leptomeningeal enhancement (red arrows, see Supplementary Table 1). (**B**) Heatmap showing pairwise correlations at single-cell transcriptome level for LUAD-LM patient CSF-CTCs, human non-small cell lung cancer cell line H358, and human lung adenocarcinoma patient-derived xenograft (PDX) cells MBT15 and PT45. (**C**) ***Top***: degree of heterogeneity among cell measured by the mean correlation coefficient within individual samples. ***Bottom***: heterogeneity analysis showing the mean correlation coefficients for CTCs within individual CSF samples (intra-patient), among CSF samples (inter-patient), and for cells within individual two PDX samples and H358 cell line (intra-others). (**D**) Heatmap of differentially expressed genes (adjusted *P*-value < 0.05) that are exclusively or preferentially expressed in one individual LUAD-LM patient. The names of selected genes are labeled.

A total of 1,300 differentially expressed genes were identified in individual CSF samples (*P*-value < 0.05, fold change >1.5; Figure 3D). Functionally important gene families that vary across individual patients include serpins (SERPIN), galectins (LGALS), claudins (CLDN), secretoglobins (SCGB) and solute carrier family (SLC). These gene family members are the major contributors of the patient-specific signatures.

### The majority of the CSF-CTCs are in the non-cycling state in LUAD-LM patients

LUAD-LM patients tend to have poor prognosis. After treatment, residual tumor cells disseminate rapidly in CSF within several months. We analyzed the cell-cycle state of the CSF-CTCs based on the single-cell transcriptomes. On average, high-cycling cells only account for 7.2% in LUAD-LM patient (4% in P1, 11% in P2, 14% in P4, 3% in P6 and 4% in P7), which is much fewer than the H358 cell line (36%) and two PDX samples (33% in MBT15, 25% in PT45) (Figure 4, A and B, and Supplemental Figure 4A).

**Figure 4.**
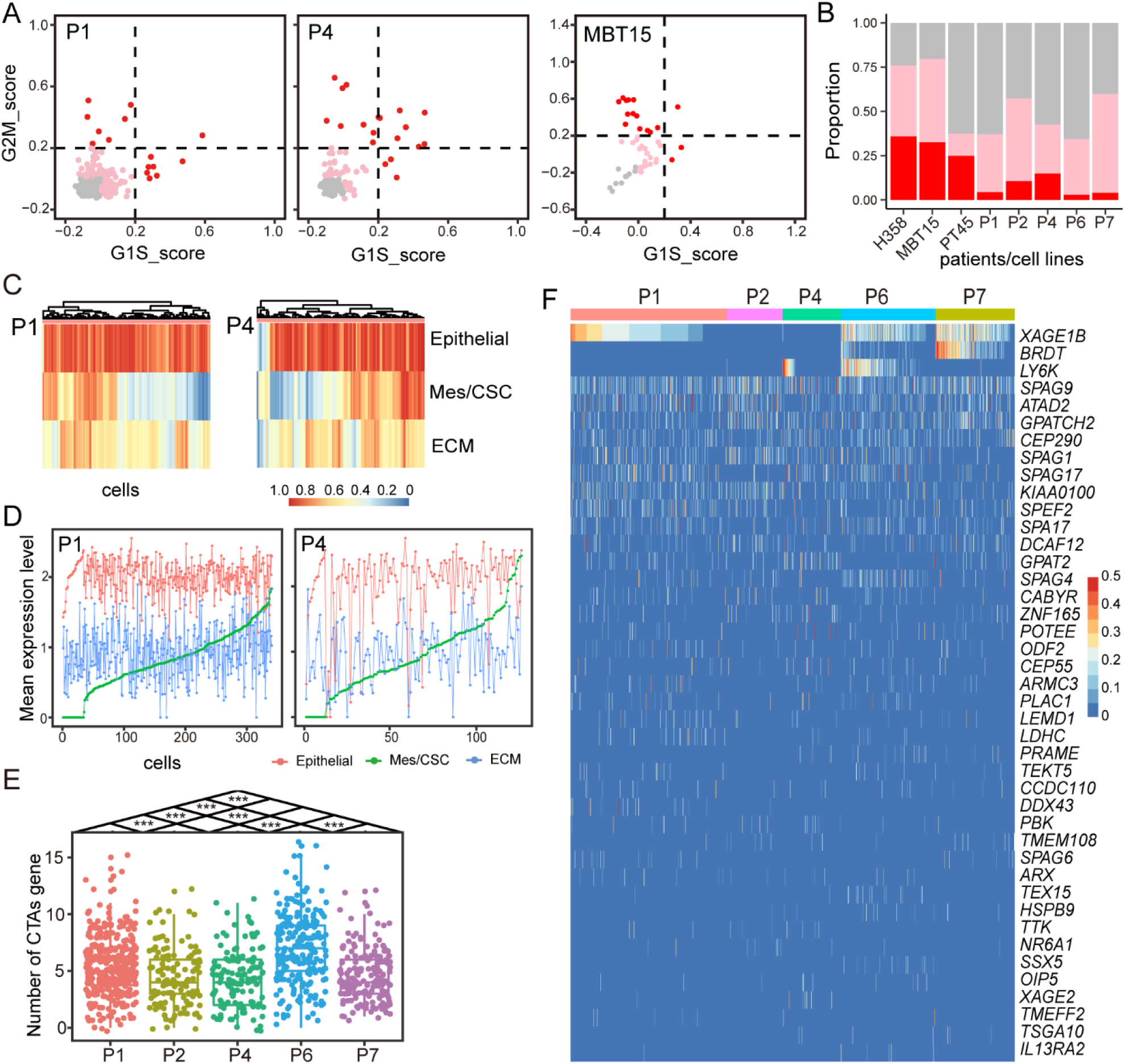
CSF-CTC gene expression profiles of cell cycle genes, cancer-testis antigens (CTAs) and genes related to partial EMT. (**A**) Cell cycle state of individual CSF-CTCs (dots) estimated based on relative expression of G1/S (*x*-axis) and G2/M (*y*-axis) genes in patient P1and P4, and MBT15. Cells are colored by inferred cell cycle states (cycling cells: score > 0.2, red; intermediate: 0 < score ≤ 0.2, pink; noncycling cells: score ≤ 0, gray). (**B**) Summary of cell cycle state proportions (*y*-axis) enriched in each sample (*x*-axis). The color code is consistent with in Figure 4A. (**C**) Unsupervised clustering using the GSVA (Gene Set Variation Analysis) score of epithelial genes, mesenchymal/CSC genes and extracellular matrix genes in P1 and P4 CSF-CTCs. Epithelial genes: *CDH1, EPCAM, KRT18, KRT19, KRT7, KRT8* and *MUC1*; mesenchymal/CSC genes: *FN1, VIM* and *CD44*; extracellular matrix genes: *LAMA3, LAMA5, LAMB2, LAMC1, ITGA3, ITGB4* and *CD47*. (**D**) Line chart of average normalized expression level (*y*-axis) of epithelial genes (red), mesenchymal/CSC genes (green) and extracellular matrix genes (blue) in P1 and P4. CSF-CTCs were ranked by average normalized expression level of mesenchymal/CSC genes (*x*-axis). (**E**) Boxplot of the number of expressed CTAs (*y*-axis) in CSF-CTCs from five patients (*x*-axis). (**F**) Heatmap of single-cell expression profiles of cancer-testis antigens in CSF-CTCs from five LUAD-LM patients.

### Cancer stemness and partial epithelial-to-mesenchymal transition (EMT) in CSF-CTCs

Cancer stem cells (CSC) have the properties of asymmetrically dividing, differentiation and self-renewal, as well as increased intrinsic resistance to therapy (29). The expression levels of lung CSC candidate biomarkers were investigated, including *PROM1*(*CD133*) (30), *CD44* (31), *ALDH1A1* (32), *ALDH1A3* (33), *ALDH3A1* (34) and *ABCG2* (35) (Supplemental Figure 4b). Elevated aldehyde dehydrogenase (ALDH) activity is a hallmark of CSC in NSCLC. Among the three ALDHs we examined, 162 (16.7%) CSF-CTCs have detectable expression of *ALDH1A*. 27.7% (268/967) CSF-CTCs across five LUAD-LM patients have CD44 expression, which is a stem cell marker for CTC aggregation and polyclonal metastases (36). *PROM1* and *ABCG2* positive CSF-CTCs are extremely rare. 57 CTCs have both CD44 and ALDH1A1 expression.

Epithelial-to-mesenchymal transition (EMT) has been suggested as a driver of epithelial tumor spreading (37). During the EMT process, epithelial cells lose cell-cell adhesion and cell polarity, in order to gain migration and invasion capabilities to behave like multipotent mesenchymal stem cells. Almost all CSF-CTCs have high expression of epithelial markers (Figure 4, C and D, and Supplemental Figure 4C). However, we discovered partial EMT process in these CSF-CTCs, which is defined as tumors cells exhibiting both mesenchymal and epithelial characteristics(38). Based on three markers (*FN1, VIM, CD44*), 113 cells in P1 (33.2%) and 54 cells in P4 (42.5%) have both epithelial and mesenchymal/CSC markers score greater than 0.5 (Figure 4, C and D), suggesting partial EMT in these patients. However, other patients only have a few cells with both epithelial and mesenchymal/CSC characteristics (6 in P2, 1 in P6 and 15 in P7) (Supplemental Figure 4C). Although these CSF-CTCs have EMT features, they lack expression of N-cadherin, classical EMT transcription factors (*ZEB1/2, TWIST1/2* and *SNAIL1/2*) or the EMT regulator TGFβ(39).

We also examined the extracellular matrix (ECM) related markers, which is another class of EMT features. Compared to normal CSF cells, the ECM receptor interaction pathway was significantly enriched in CSF-CTCs (FDR < 0.05, Figure 2E). We selected core enrichment genes of ECM receptor interaction pathway, including laminins (*LAMA3, LAMA5, LAMB2, LAMC1*) (40), integrins (*ITGA3, ITGB4*) (41) and *CD47* (42). Abundant expression of ECM genes is observed in all patients, which could be a common feature of CSF-CTCs (Figure 4c, d and Supplemental Figure 4C). These results suggest that the upregulation of ECM related genes may contribute to the generation of CTCs from solid tumor sites or to the survival of cancer cells deprived of microenvironmental signals as they circulate in the CSF.

### Cancer-testis antigens (CTAs) in CSF-CTCs contribute to the among patient heterogeneity

Tumor cells frequently express cancer-testis antigens (CTAs) whose expression is typically restricted to germ cells, providing unprecedented opportunities for clinical development of cancer diagnosis and immunotherapy(43). A recent study has demonstrated the extensive heterogeneity of CTAs in LUAD single-cell data (PDXs and cell lines) (44). However, little is known about the heterogeneity of all possible CTAs expressed in CSF-CTCs from LUAD origin. We examined the expression of 276 selected CTAs (http://www.cta.lncc.br/modelo.php) in CSF-CTCs. We discovered that patients P1 and P4 CTCs have significantly larger numbers of expressed CTAs than other patients CTCs, and substantial inter-tumor heterogeneity and intra-tumor heterogeneity of CTAs among CTCs (Figure 4, E and F). Expression of *XAGE1B* is only observed in P1, P6 and P7, whereas *BRDT* expression is restricted in P6 and P7. *LY6K* is specific to subset of CTCs in P4 and P6. *SPAG9* is ubiquitously expressed in 41.3% of CTCs in all five patients (399/967) at high level, with the potential to serve as a target for immunotherapy (45).

### Characterization of a case of cancer of unknown primary site (CUP) through CSF-CTC single-cell transcriptomes

Patient P8, a 49-year-old male, was diagnosed with cancer of unknown primary site (CUP) in 2017. CUP is a well-recognized clinical disorder, accounting for 3-5% of all malignant epithelial tumors. Metastatic adenocarcinoma is the most common CUP histopathology (80%)(46). P8 showed multiple metastases including cervical lymph nodes and LM (Figure 5A). The immunohistochemistry (IHC) results of left lymph nodes biopsy indicated that the metastatic adenocarcinoma with positive epithelial markers (CK pan, CK7, CK8, CK18, CK19 and MUC1; Figure 5, B and C) and a prolactin-induced protein PIP/GCDFP15 (Figure 5D, upper panel), which is a small secreted glycoprotein whose expression is generally restricted to cells with apocrine properties (47). The proliferation marker MKI67 was partially positive (Figure 5D, lower panel). Therefore, the primary tumor was epithelial origin with apocrine properties. Based on the IHC results, we could exclude the high possibility of the following locations of the primary tumor: lung cancer (markers NAPSA−, TTF1−, P63−, synaptophysin/SYP−; Figure 5E) (48, 49), gastrointestinal cancer (VIL1/Villin−; Figure 5F) (50), prostate cancer (KLK3/PAS−; Figure 5G)(51) and liver cancer (GPC3−; Figure 5H)(52). P8 had partial response (PR) to chemotherapy (paclitaxel liposome for injection and cisplatin), but the disease had recurred with LM after the treatment (Figure 5A).

**Figure 5.**
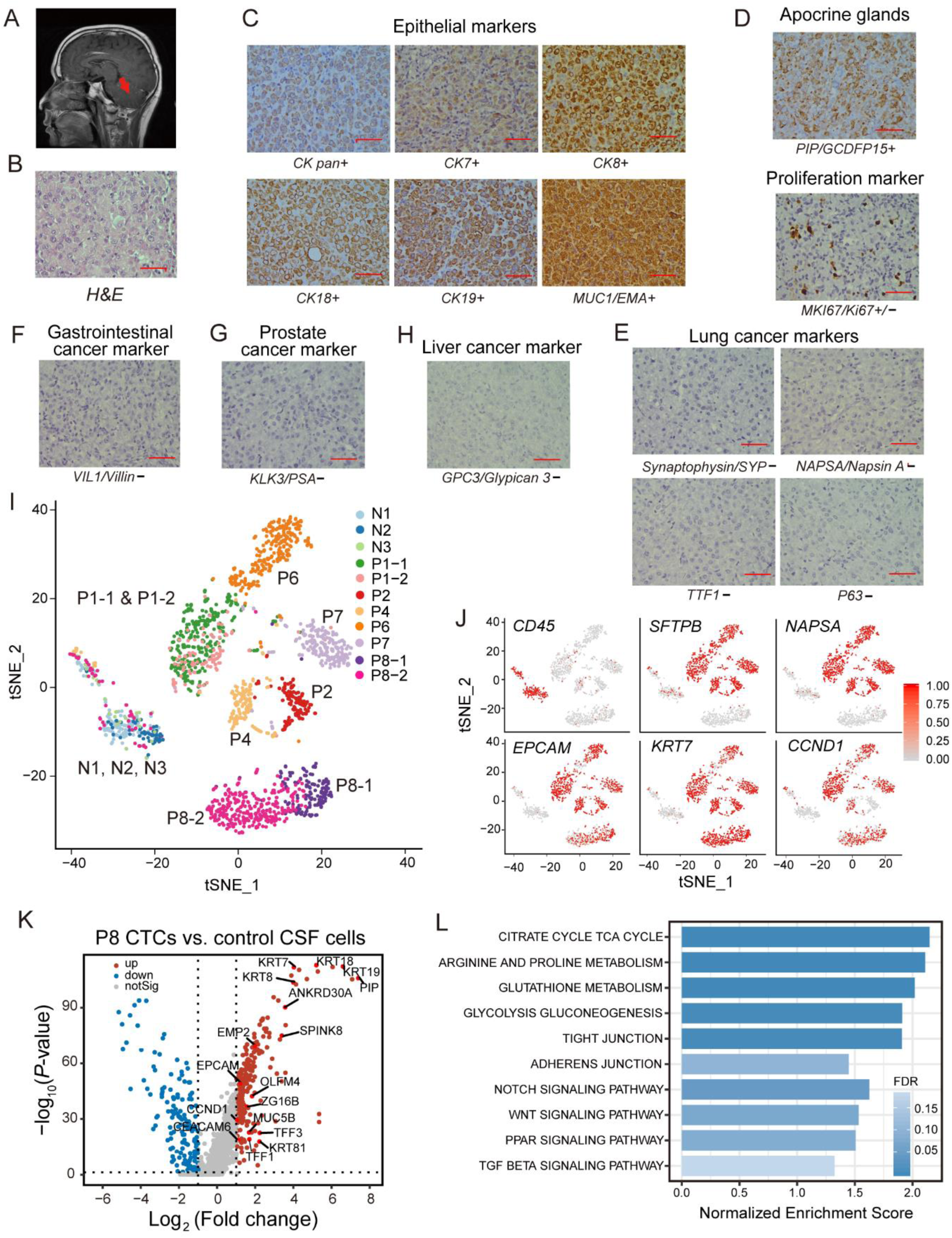
Investigation of a leptomeningeal metastases (LM) case of cancer of unknown primary site (CUP) through scRNA-seq profiling of CSF-CTCs. (**A**) Brain MRI results of patient P8, demonstrating leptomeningeal enhancement in cerebellum (red arrows). (**B-H**) H-E staining (***B***) and immunohistochemistry (IHC) of left lymph nodes biopsy for epithelial markers CK pan+, CK7+, CK8+, CK18+, CK19+, MUC1+ (***C***), apocrine gland marker PIP/GCDFP15+ and proliferation marker MKI67/KI67+/− (***D***), lung cancer markers NAPSA−, TTF1−, P63−, synaptophysin/SYP− (***E***), gastrointestinal cancer marker VIL1/Villin− (***F***), prostate cancer marker KLK3/PAS− (***G***), and live cancer marker GPC3−/Glypican− (***H***) (+: positive, −: negative, +/−: partly positive, scale bar represents 200 μm). **i**, t-SNE plot of gene expression clustering of patient P8 with normal CSF samples (N1, N2 and N3) and LUAD-LM CSF samples (P1, P2, P4, P6 and P7). P8-1 and P8-2 are two independent CSF samples 6 months apart. (**J**) Feature plots demonstrating expression of selected genes in the t-SNE plot (Figure 5A). Scaled expression levels are depicted using a red gradient. *CD45*: leukocyte marker. *SFTPB* and *NAPSA*: lung markers. *EPCAM* and *KRT7*: epithelial marker. *CCND1*: proliferating cells marker. (**K**) Volcano plot of significant differential gene expression (adjusted *P*-value < 0.05; fold-change > 2) between P8 CSF-CTCs and leukocytes. (**L**) Significantly enriched KEGG pathways by GSEA analysis in P8 CSF-CTCs compared to leukocytes (FDR: 0.05-0.20).

In the scRNA-seq profile with other LM patients (P1-P7), P8 CSF-CTCs formed a single cluster on the t-SNE clustering plots, independent from LUAD-LM CTCs and normal CSF cells (Figure 5i). There is some degree of separation within this cluster between samples P8-1 and P8-2, which were collected with a six-month time interval, reflecting disease progression (Figure 5I). P8 CSF-CTCs were defined by the epithelial signature and lack of CD45 expression (Figure 5J). Consistent with the immunohistochemistry results (Figure 5E), none of the lung origin markers were expressed. In addition, upregulated genes in LUAD-LM CTCs (for example, *MMP7, SCGB3A2, C3, CDH1* and *EGFR*) are not detected in P8, except for *CEACAM6* which is shared across P8 and all LUAD-LM patients (Figure 5K). The gene set enrichment analysis (GSEA) revealed active metabolism and tight junction pathway (FDR < 0.05) as the characteristics of P8 CTCs (Figure 5L).

Based on the single-cell transcriptome profiles, 40 P8 cluster defining genes (Figure 6, A and B) were selected from the DEG list between P8 and LUAD-LM CSF-CTCs based on criteria listed in Supplementary Table 5. Among those genes, the top two candidates are *PIP* and *ANKRD30A. PIP* is a cytoplasmic marker commonly used to identify breast cancer, but not exclusively, as its expression was also found in several other types of human cancers including prostate, sweat and salivary gland cancer (47). *ANKRD30A* is restricted to normal breast, normal testis, normal prostate and also detected in breast cancer as a breast cancer specific marker (53) and in prostate cancer (54) (Figure 6, A and B).

**Figure 6.**
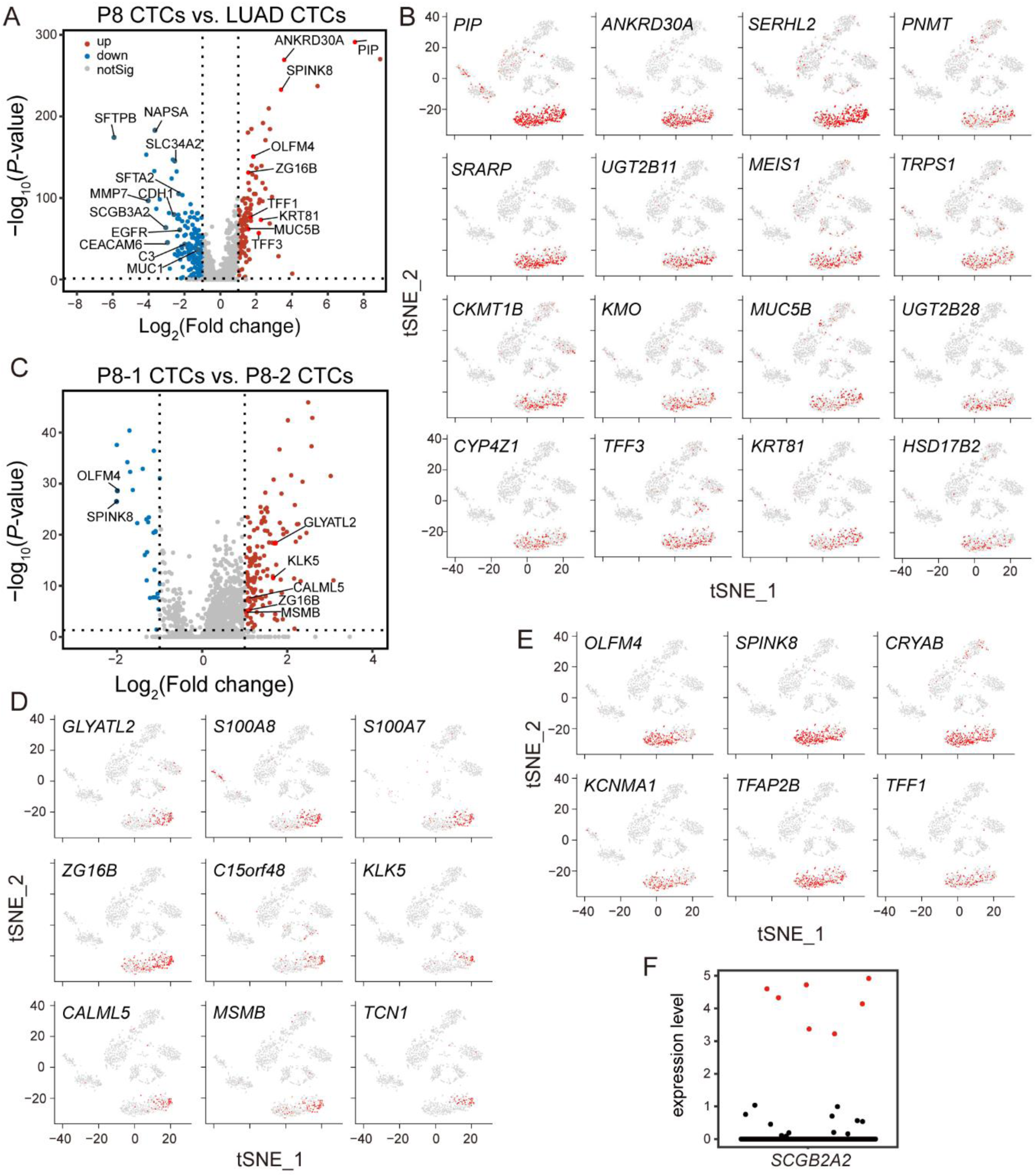
The gene expression profile and characterization of CSF-CTCs in P8-1 and P8-2. (**A**) Volcano plot of CSF-CTC differential gene expression (adjust *P*-value < 0.05) between P8 and LUAD-LM patients. Up-regulated (*up*) and down-regulated (*down*) genes are defined using a fold-change cutoff of 2. Selected gene names are labeled. (**B**) Feature plots of 16 top P8-specific genes (see selection criteria in Supplementary Table 5) on the t-SNE plot (Figure 5I). Scaled expression levels are depicted using a red gradient (grey denotes lack of expression). (**C**) Volcano plot of CSF-CTC differential gene expression (adjust *P*-value < 0.05) between P8-1 s and P8-2. Up-regulated (*up*) and down-regulated (*down*) genes are defined using a fold-change cutoff of 2. Selected gene names are labeled. (**D-E**) Feature plots of P8 stage-biased genes (see selection criteria in Supplementary Table 6) on the t-SNE plot (Figure 5I). Scaled expression levels are depicted using a red gradient (grey denotes lack of expression). Nine P8-1 biased genes (***D***) and six P8-2 biased genes (***E***) were plotted. (**F**) Normalized expression levels of *SCGB2A2* in all cells (Figure 5I). Seven cells in P8 with high expression of *SCGB2A2* are labeled in red.

202 genes were differentially expressed between P8-1 and P8-2 (Figure 6C). 9 genes are preferentially expressed in P8-1 (Figure 6D, Supplementary Table 6), whose expression could not be detected in most CTCs at the later stage P8-2. *OLFM4, SPINK8, CRYAB, KCNMA1, TFAP2B* and *TFF1* are significantly upregulated during tumor progression in P8-2 CTCs (Figure 6E, Supplementary Table 6).

Seven P8 CTCs have high expression of the *SCGB2A2* genes (Figure 6F), a carcinoma marker of breast origin including primary tissues, metastatic tissues and blood-CTCs (55, 56). *SCGB2A2* is also positive in some tissues of gynecologic malignancies (57), but P8 is a male. In addition, *SCGB2A2* is also associated with salivary gland cancer (58).

The proliferation ability of P8 CTCs is similar to LUAD-LM CSF-CTCs, with 3/131 (P8-1) and 11/262 (P8-2) cells in high-cycling state (Supplemental Figure 6A). Interestingly, a large proportion of (118/393) P8 cells have *PROM1* (*CD133*, a classic CSC marker) expression, which is different from LUAD CSF-CTCs (20/967). The expression levels of epithelial, mesenchymal and CSC genes were also examined. All P8 CTCs have high expression of epithelial genes. CTCs (290/393) with stem-like phenotype (*PROM1+CD44*) are in epithelial state(59) and about 10% (36/393) cells have detectable expression of mesenchymal genes (*FN1+VIM*) (Supplemental Figure 6B).

## Methods

### Patients information and sample collection

All human sample materials used in this research are collected at Huashan Hospital, Fudan University. The consent forms and the proposed studies were approved by Ethics Committee. Clinical information of patients was listed in Supplementary Table 1.

### Cell Sorting and single-cell preparation

Antibody (CD45, CD3, CD19, BD Biosciences) and labeling dye for live cells (Calcein Blue AM, Life Technologies, CA) were used per manufacturer recommendations. Live cells (Calcein Blue AM+) in normal CSF samples (N: N1-N3) and live tumor cells (Calcein Blue AM+, CD45−) in patient CSF samples (P: P1-P8) were selected for sequencing (Figure 1A, and Supplementary Table 1). Among all of the CSF samples, 3,792 single cells were selected for sequencing (N: 624 cells; P: 3,168 cells; Table S1). 168 blood-T cells (Calcein Blue AM+, CD45+, CD3+) and 168 blood-B cells (Calcein Blue AM+, CD45+, CD19+) were also sorted for sequencing (Supplementary Table 1).

Target cells were sorted into pre-prepared 96-well plates by FACS (fluorescence-activated cell sorting). Single-cell lysates were sealed, vortexed, spun down, placed on dry ice and transferred immediately for storage at −80°C.

### SMRAT-seq2 library construction and sequencing

Library for isolated single cell was generated by SMART-Seq2 method (60) with the following modifications: RNA were reverse transcribed with Maxima H Minus Reverse Transcriptase (Thermo Fisher Scientific, MA), and whole transcriptome were amplification using KAPA HiFi Hot Start Ready Mix (KAPA Biosystems, MA). cDNA library was purified using Agencourt XP DNA beads (Beckman Coulter, CA) and quantified with a high sensitivity dsDNA Quant Kit (Life Technologies, CA). It is worth mentioning that full length cDNA libraries were tagmented, and then only 3’ end sequence (500-1000bp) was amplified and enriched for sequencing on an Illumina HiSeqX machine, which is different from traditional SMART-Seq2 method of full tagmented-libraries sequence.

### scRNA-seq expression analysis

The Illumina sequencing data were demultiplexed based on sample barcodes. Adapter sequences, poly T and residue barcodes were trimmed using custom scripts. We used UMI-tools (61) to remove UMIs and trim-galore (62) to remove low-quality bases. The filtered reads were aligned to the human reference genome (hg19) by STAR (63) and BAM files were prepared by SAMtools (64). Gene expression counts were obtained by FeatureCounts (65). Fastqc (66) and multiqc (67) were used for quality control and reports during each step.

Genes expressed in less than 10 cells were filtered out from gene expression matrix of CSF samples. Individual cells with fewer than 600 covered genes and over 20% mitochondrial reads were filtered out, and 1,986 single cells remained (401 immune cells and 1,585 CTCs) for subsequent analysis using the Seurat 3.0 software package(68, 69) (Table 1).The mean number of genes detected per cell is 830 for immune cells and 1,870 for tumor cells, respectively.

When we analyze the transcriptome characteristics of CTCs, we selected tumor cells with more than 1000 covered genes and have 1360 CTCs retained (340/ P1, 122/P2, 127/P4, 206/P6, 172/P7, 393/P8) for analysis. The mean number of genes detected per LUAD CSF-CTCs was 2070.

### Clustering and marker expression analysis for cell type identification

Cells were clustered by non-supervised t-SNE dimensionality reduction (70) based on their gene expression counts. The cells were separated into groups with indication of cryptic inner-group connection. The cluster-specific marker genes were identified by the FindAllMarkers function in Seurat 3.0. Single-cell RNA-Seq data of two human lung adenocarcinoma patient-derived xenograft (PDX) samples (LC-PT-45, PT45; LC-MBT-15, MBT15) and human non-small cell lung cancer cell line (H358 cell line) were obtained from NCBI Sequence Read Archive with accession number GSE69405 (28). These data were filtered using the same pipeline as CSF-CTCs and only cells with more than 1,000 genes were included for analysis.

To infer the cell type identity, Seurat 3.0 was used to generate expression heatmaps of selected gene markers of known cell types, including T cells (*CD2, CD3D, CD3E* and *CD3G*), B cells (*CD19, MS4A1, CD79A* and *CD79B*), monocyte (*CD14, CD68* and *CD163*), lung cells (*SFTPA1, SFTPA2, SFTPB* and *NAPSA*), epithelial cells (*EPCAM, CDH1, KRT7, KRT8, KRT18* and *MUC1*), proliferative cells (*CCND1* and *TOP2A*) (71-77). The ImmuneScore was computed based on ESTIMATE (Estimation of STromal and Immune cells in MAlignant Tumors using Expression data) R package(17).

### Differential gene expression and pathway enrichment analysis

Significantly differential expression gene (DEGs) between samples were detected by DESeq2(78) using normalized gene expression counts, at a adjust p-value cutoff of 0.05 and a fold-change cutoff of 2. GSEA (gene set enrichment analysis) (79, 80) were used for functional enrichment analysis of KEGG pathways (80-83).

### Cell cycle analysis

Cell cycle assignment was performed in R version 3.6.0 using the CellCycleScoring function included Seurat 3.0 package. Cycling cells are defined as cells with either G1/S.Score or G2/M.Score greater than 0.2. The criterion of intermediate cells is 0 < G1/S.Score or G2/M.Score ≤0.2. The rest cells are defined as non-cycling cells.

## Discussion

### The clinic value of CSF samples in lung cancer brain metastases

The majority of LM is from solid malignancies of primary breast and lung cancers (84) and LM remains a clinical challenge. The cerebrospinal fluid (CSF) is in direct contact with tumor cells in LM, therefore liquid biopsy of CSF will reflect the real-time status of LM. CSF cfDNA (cell-free DNA) can be used to characterize and monitor the development of NSCLC-LM, and mutations detected in the cfDNA will inform the clinical therapy (85). However, a recent research revealed 53.2% cfDNA mutations in blood of cancer patients had features consistent with clonal hematopoiesis which were not real mutations of tumors (86), and the inability to trace the tumor cells limit the application of cell-free method. It has been shown that CTC counts in CSF by CellSearch is associated with clinic prognosis of lung cancer LM (12), but further studies were needed to fully characterize the CSF-CTCs in LUDA-LM patients. To fill this critical gap, our study serves as the first single-cell transcriptome profiles of LUAD CSF-CTCs by scRNA-seq in multiple patients. We established a protocol for successful isolation of CTCs for scRNA-seq in CSF samples. The CSF-CTC population is enriched for meta-static precursors, and their single-cell transcriptome signatures will facilitate early detection of LUAD-LM and the identification of potential therapeutic targets.

### Single-cell transcriptome signatures defining the CSF-CTCs in lung adenocarcinoma leptomeningeal metastases (LUAD-LM)

To define the LUAD-LM CSF-CTCs, we discovered a number of important genes significantly higher expressed in patient CSF-CTCs compared to normal CSF cells including several lung cancer markers and proliferation makers. For example, the expression of cycle gene *CCND1* in 61.7% (596/967) CSF-CTCs was much broader and higher than MKI67 in 12.5% (121/967) CSF-CTCs (Figure 2, B and D). Therefore, *CCND1* is more suitable to serve as proliferation marker for LUAD CSF-CTCs diagnosis though MKI67 is a classical proliferation marker which is commonly used in clinic immunohistochemistry. *SLC34A2* is another commonly expressed gene in CSF-CTCs (Figure 2D). Its expression was only found in the apical membrane of type II alveolar epithelium cells (ATII) (87, 88), providing the potential as a candidate LUAD CSF-CTC specific marker. In addition, the high expression of EGFR, ERBB2 and EMP2 also correspond to the characteristics of LUAD cells (Figure 2D). EGFR and ERBB2 are members of epidermal growth factor (EGF) receptor family of receptor tyrosine kinases, and have been implicated in the development of many cancers. The genetic changes of EGFR and ERBB2 lead to receptor overexpression and promote constitutive receptor activation, a process that stimulates cancer development (89). Epithelial membrane protein 2 (EMP2) is by far most highly expressed in AT1 cells of lung and serves as a platform for integrin signaling, supporting cell adhesion to extracellular matrix (ECM) and other cytoskeletal functions (90, 91).

Several previously known markers genes are not expressed in the CSF-CTCs we examined. *TTF1* is a major transcription factor regulating genes expression in airway epithelial cells, including surfactant proteins *SFTPA* (92), *SPTPB* (93), *SFTPC* (94) and *SCGB1A1* (CC10) (95). *TTF1* has been used for clinical diagnosis of human LUAD (96, 97). However, its expression in CSF-CTCs was not upregulated compared to immune cells which limited its application in LUAD-CTCs identification. Secretoglobin superfamily genes are diagnostic markers for cancers, and the most studied member *SCGB1A1* (CC10) is considered as a tumor suppressor and expressed in less than 10% of human NSCLCs (98). In contrast, *SCGB3A2* is a useful immunohistochemical marker for NSCLCs, particularly adenocarcinomas (19). *SCGB1A1* is not expressed in CSF-CTCs, whereas SCGB3A2 are broadly and highly expressed in the majority of the cells, which could serve as a novel marker for LUAD CSF-CTCs (Figure 2D).

### Energy metabolism and cell adhesion pathways are significantly enriched in LUDA CSF-CTCs

Glucose is the main energy metabolite and it is transported through endothelial cell glucose transporter GLUT-1 across blood-brain barrier to meet the high energy demands of the brain tissue (99). A previous study using experimental brain metastases models has shown that the utilization of glucose was enhanced in both glycolytic and pentose phosphate pathways (100). This is also observed in patient CSF-CTCs-with glucose-associated pathways significantly up-regulated (Figure 2E). Tumor cells also have active and rich amino acid metabolism to ensure the sufficient protein synthesis for tumor survival.

The adhesion between CTCs and endothelial cells is essential in tumor metastasis (101), by facilitating the circulation and invasion of the metastatic cells (102). The up-regulation of adhesion-related genes in CSF-CTCs are also important for CTCs clusters formation, which have been detected by CellSearch technology previously (12). Blood-CTC cluster termed “circulating tumor microemboli” (CTM), or CTC-WBC cluster have been described with higher malignancy and have a better change to survive in bloodstream (103, 104). Plakoglobin (JUN) was identified as a key mediator of tumor cells clustering in human breast CTCs, and we detected the expression of plakoglobin in a subset of CSF-CTCs (103). However, the critical gene for CTC-neutrophil cluster formation, *VCAM1*, is not detected in our CSF-CTCs (104). CSF-CTC cell adhesion characteristics could enhance the CSF tumor cluster formation and the migration of CTCs, which is a potential target for tumor therapy strategies.

### Heterogeneity of LUAD-LM tumor cells in CSF samples

#### Spatial and temporal heterogeneity

The LM locations in the brain are quite diverse in all patients examined. Adaptation to different local brain environment may contribute to the gene expression heterogeneity of CSF-CTCs among patients. The scRNA-seq approach could also detect potential temporal heterogeneity during tumor progression. We obtained CSF samples from two different time points for both patient P1 (P1-1 vs. P1-2) and P8 (P8-1 vs. P8-2). Collected within a 2-month time interval, P1-1 and P1-2 have similar single-cell expression profiles and CSF-CTCs from both datasets form a single homogeneous cluster (Figure 2A), indicating similar transcriptome patterns. In contrast, P8-1 and P8-2 were collected 6-month time apart as the patient’s condition worsened. Although they are still in the same cluster, we observed clear separation of single-cell expression profiles on the t-SNE plot, which may correspond to the transcriptome signatures of LM progression (Figure 5I). Systematic sampling over a time course in future studies will allow better characterization of the spatial and temporal heterogeneity.

#### Gene expression basis of CTC heterogeneity

One major advantage of single-cell RNA-seq approach is the ability to characterize the expression variation among individual cells. We found significant greater expression heterogeneity among patient CSF samples than among cells within patient samples, suggesting the need of personalized diagnosis and expression profiling. Differential gene expression analysis revealed the following gene families vary across patient CSF-CTC samples (Figure 3D). Serpins are highly expressed in lung cancer brain metastatic cells, promoting tumor cell survival and metastases through the inhibition of plasmin generation (105). Galectins mediate cell-cell and cell-ECM interactions and modulate cell signaling and the tumor microenvironment, which could influence the tumor invasion, migration, metastasis and progression (106-108). Claudins (CLDN) are a family of integral membrane proteins located in epithelial cells in tight junctions bearing tissues. Their expression was found in many lung cancer histological types, and overexpression could results in tumor spreading (109). In addition, S100 family were highly expressed in P6, whereas P7 has elevated expression of many ribosomal protein genes (RPL and RPS). These different genes and gene families all contribute to the heterogeneity of LUAD-LM tumor cells.

#### Mutational profile heterogeneity

In addition to tumor location in leptomeningeal, the mutational profiles of tumor cells may also contribute to the heterogeneity. Mutations in cerebrospinal fluid cell-free DNA (CSF cfDNA) were detected by next-generation sequencing (Supplementary Table 4). The discovery of common driver mutations and the development of targeted therapies dramatically improved the intracranial efficacy resulting in prolonged survival. Activating mutations of the epidermal growth factor receptor (EGFR) and anaplastic lymphoma kinase (ALK) rearrangements are key in the development of brain metastases(110, 111). P1 belongs to this category with EGFR (19del) and ALK (Arg1192Trp) mutation, along with TP53 (Trp53Ter) mutation and low frequency of KRAS mutation (Supplementary Table 4). EGFR (Leu858Arg) and TP53 (Arg248Gln) mutations were found in P4, and P7 only has TP53 (Asp281Tyr) mutation. We have not performed mutation detection of CSF cfDNA by NGS -in P2 or P5. These different mutational profiles, together with different treatment strategy, also contribute to the greater among patient heterogeneity.

#### Potential epigenetic heterogeneity

*SCGB3A1*, a tumor suppressor in many cancer types, loses expression due to promoter methylation (112). Interestingly, *SCGB3A1* was only highly expressed in CSF-CTCs of P2, P6, P8, but not in other LUAD-LM patients, indicating potential methylation heterogeneity among cells and patients.

### CSF immune function and cancer

Normal CSF environment has its own specialized immune cell composition, including T cells and Monocytes but not B cells (113-115). Our scRNA-seq results of control patient samples confirmed that composition at single-cell transcriptome level. When tumor cells infiltrate CSF, the central immune system is activated to combat the tumor cells. Efficacy of cancer immunotherapies is partly depending on the amount and properties of tumor infiltrating lymphocytes (116). A recent single-cell RNA study showed great heterogeneity within the tumor regulatory T cells (Tregs) and tumor-infiltrating CD8+ T cells, and a low ratio of “pre-exhausted” to exhausted T cells was related to worse prognosis of lung adenocarcinoma (117). In addition, For patients with HIV infection, there are rare CNS immune cell subsets in CSF that may perpetuate neuronal injury (15). Therefore, analyzing the immune cell composition in pathological CSF samples would be informative for understanding the interactions between CSF microenvironment and metastatic cancer cells. The tumor infiltrating immune cells in CSF samples is worth future research.

### The metastatic process – partial EMT and stemness

EMT is a process related to tumor invasion and metastases. It has been reported that some NSCLC blood-CTCs have a dual epithelial-mesenchymal phenotype (118). Similarly, we discovered abundant expression of epithelial genes in LUAD-LM CSF-CTCs, and a small subset of CSF-CTCs expressed mesenchymal genes (*VIM* and *FN1*) but without the expression of EMT transcription factors which are necessary for EMT. However, cells with high expression of mesenchymal genes and low expression of epithelial genes are extremely rare in LUAD-LM CSF-CTCs (only 2 cells in Supplemental Figure 4D) which is a major difference with NSCLC blood-CTCs.

Notably, we also identified the unexpected abundant expression of extracellular matrix (ECM) genes in CSF-CTCs which consistent with ECM characteristics of blood-CTCs in pancreatic, breast, and prostate origin (119). Tumor stroma-derived ECM signaling plays an important role in targeting cancer cell metastasis (120). The cell-autonomous expression of ECM genes in CSF-CTCs may contribute to the dissemination of cancer. CD44 as the important stem cell marker of CSF-CTCs improved tumor initiation capacities of CTCs. We did not observe any correlation between CD44 expression and enrichment for the mesenchymal genes (*VIM* or *FN1*) within single CSF-CTC, suggesting that stem cell and EMT markers are not intrinsically linked in CSF-CTCs (Supplemental Figure 4E) which is also observed in pancreatic blood-CTCs (119). The advancement of the CSF-CTC metastatic characteristics and comparison with NSCLC blood-CTCs will provide a much better understanding of the mechanisms of LUAD-LM.

### The power of single-cell RNA sequencing in CSF samples for the diagnosis of CUP origin

CUP is a well-recognized clinical disorder, which accounts for 3-5% of all malignant epithelial tumors with poor prognosis due to treatment of a non-selective empirical therapy. Identification of the primary tumor type will greatly inform the treatment strategies, but it is extremely challenging. It is estimated that about 80% CUP cases are metastatic adenocarcinoma (46). Our studies also focused on this most common CUP by enrolling one CUP patient of metastatic adenocarcinoma (P8).

In order to pinpoint the organ and tissue of origin, patient history, physical examination, serum markers, histological data and state-of-the-art imaging results have been examined and the primary origin remains inconclusive. For CUP patients with leptomeningeal metastases only, the CSF-CTCs are the available tissue sample for the diagnosis of the primary origin of CUP. Since P8 is a CUP case with multi-site metastases, we have the biopsy of left lymph nodes to perform immunohistochemistry (IHC) to pair with our single-cell RNA-seq data. The scRNA-seq data in CSF-CTCs and IHC results in lymph nodes revealed the epithelial origin and it is less likely to have lung, prostate, gastrointestinal or liver origin, providing crucial diagnostic information for this patient. The cluster-defined genes, *PIP* and *ANKRD30A*, are exclusively expressed in P8 compared to other LUAD-LM CSF samples, indicating sufficient evidence to diagnose the primary site as breast cancer, sweat/salivary gland cancer and prostate cancer,-Interestingly, when we checked the expression of SCGB2A2, a classical marker of breast cancer, 7 CTCs from P8 have high expression levels (Figure 6F) whereas other CSF-CTCs have little to no expression. This scRNA-seq result enhanced the diagnosis directions of breast cancer or sweat/salivary gland cancer origin. Then, further investigation has been made on the possibilities of breast, prostate and sweat/salivary gland cancer, but no evidence has been found despite extensive imaging examinations. The definitive conclusion could not be made because the patient passed away and refused autopsy.

Immunohistochemical markers are the most important diagnostic tools in establishing tissue origin of CUP, but scRNA-seq has far better sensitivity, especially for CSF samples with low tumor burden. The successful detection of 7 cells expressing *SCGB2A2* in patient P8 is one example of the advantage of single-cell sequencing over bulk RNA-seq or IHC. As the first CUP case with scRNA-seq data in CSF-CTCs, we were able to achieve a comprehensive characterization of the transcriptome pattern in every P8 tumor cell, as well as the discovery of potential biomarker expressed at a low frequency in specific cells. With continuous advancement of scRNA-seq technology and deceasing sequencing cost, additional scRNA-seq datasets will be available for breast cancer, sweat/salivary gland cancer, and prostate cancer. In the near future, we will be able to build transcriptome databases of multiple CUP cases and provide speedy and accurate diagnosis for CUP origin to benefit the cancer patients.

### The transcriptome signature of CSF-CTCs in the CUP case

Traditional diagnostic methods rely on CSF cytology and the examination of known marker genes. Our scRNA-seq approach can not only profile the classic markers with great sensitivity and precision, but also discover novel markers which are not well-known for cancer diagnosis. The CSF-CTCs of P8 showed low-proliferative and high-epithelial signatures as LUAD-LM patients. Unlike LUAD-LM CTCs, we found that CSF-CTCs were enriched for the stem-cell-associated gene *PROM1* and did not significantly upregulate ECM receptor interaction pathway.

Traditional diagnostic methods rely on CSF cytology and the examination of known marker genes. The scRNA-seq approach can not only profile the classic markers with great sensitivity and precision, but also discover novel markers which are commonly used for cancer diagnosis. The CSF-CTCs of P8 showed low-proliferative and high-epithelial signatures as LUAD-LM patients. Unlike LUAD-LM CTCs, we found that CSF-CTCs were enriched for the stem-cell-associated gene *PROM1* and did not significantly upregulate ECM receptor interaction pathway.

A number of genes associated with metastatic adenocarcinoma have shown to be exclusive expressed in P8 compared to LUAD-LM patients (Supplementary Table 5). *TFF1* (Figure 6E) and *TFF3* (Figure 6B) are members of the trefoil factor (TFF) family which are distributed in numerous tissues and typical constituents of mucous epithelia (121). TFFs play important roles in mucosal protection and repair, and are typically cosecreted with mucins(121). *TFF1* and *TFF3* have been reported to interact with *MUC5AC* in gastric and ocular (122). In P8 CTCs, co-expression of *TFF1, TFF3, MUC5AC* and *MUC5B* (Figure 6B) indicated the possible interaction between them.

The two P8 CSF-CTC samples collected at different tumor stages provided us a rich dataset to understand the gene expression signatures of disease progression. Among the nine genes with biased expression in the earlier stage P8-1 (Supplementary Table 6), *MSMB* (microseminoprotein beta, Figure 6d) is one of the most abundant proteins in semen(123-125) and it has been used for diagnosis of prostate cancer (126). *ZG16B*/PAUF (pancreatic adenocarcinoma-upregulated factor, Figure 6D) is a secreted protein with crucial role in pancreatic ductal adenocarcinoma (PDAC) (127-130), as well as other cancer types including epithelial ovarian cancer (131), cervical carcinoma (132), colorectal cancer (133) and oral squamous cell carcinoma (134). *KLK5* (Kallikrein-related peptidases 5, Figure 6D) is in the same gene family as the prostate-specific antigen *KLK3*, exerts key modulatory effects on many cancers(135-139) and it is selectively inhibited by SPINK, a family of serine peptidase inhibitor kazal type, which bind to their target serine proteases and inhibit their proteolytic function(140-145). Interestingly, we discovered the higher percentage of *SPINK8* (Figure 6E) expression in P8-2 CTCs compared to P8-1 CTCs, suggesting potential interaction between *KLK5* and *SPINK8* during disease progression. Like *SPINK8, OLFM4* (Olfactomedin-4, Figure 6E) is another gene with preferential expression in P8-2 CTCs. *OLFM4* has important roles in various tumors, especially in gastrointestinal malignancies with primary function as an antiapoptotic factor (146, 147). *OLFM4* is relevant to many cellular processes, including cell adhesion(148), proliferation and metastasis (149, 150), and stem cell self-renewal (151).

## Data Availability

The RNA sequencing data will be available after publication on a peer-reviewed scientific journal.

## Acknowledgements

This work is supported by the National Key Research and Development Program of China (Grant No. 2017YFA0103902), the National Natural Science Foundation of China (Grant No. 31771283), the Fundamental Research Funds for the Central Universities (Grant No. 22120190210), the Innovation Group Project of Shanghai Municipal Health Commission (2019CXJQ03) and an Auburn University Research Initiative in Cancer (ARUIC) Research Grant. M.G. is supported by the Program for Shanghai Municipal Leading Talent (2015). X.W. is supported by the USDA National Institute of Food and Agriculture (Hatch project 1018100) and a generous laboratory start-up fund from Auburn University College of Veterinary Medicine. Y.Z. is supported by Auburn University Presidential Graduate Research Fellowship.

